# Prescription intervals of medications for chronic use: a cohort study

**DOI:** 10.64898/2026.06.08.26355164

**Authors:** Ryan Muddiman, Padraic Donoghue, Jessica Gómez Lemus, Ann Sinéad Doherty, Fiona Boland, Caroline McCarthy, Frank Moriarty

## Abstract

**Purpose:** In deprescribing studies, a prescription-free gap is typically used to determine if patients discontinued their treatment. An appropriate gap depends on the typical time between prescriptions during continued use. This work aims to characterise the interval between prescriptions of chronic drugs using different methods for a cohort of older people in primary care in Ireland.

**Methods:** The empirical prescription interval was analysed for 38,154 patients for the twenty most common drug classes and the association between covariates and the interval was analysed using a multi-level model. Estimates were also compared to those obtained from the parametric waiting time distribution (pWTD) approach.

**Results:** Available covariates had consistent relationships with prescription intervals across drug classes. For example, each additional prescription issue was associated with an increase in the interval by 5.0 (NSAIDs) to 19.7 days (“Other antidepressants”). Full public health cover was associated with a -29.0 day (inhaled adrenergics) to -11.0 day (opioids) change relative to partial cover, while other/private cover had a -17.9 day (benzodiazepines and associated drugs) to -7.1 day (SSRI and SNRIs) change relative to partial cover. The pWTD also produced consistent estimates of the population interval for most drugs.

**Conclusions:** The interval varied substantially within drug classes, due to a mixture of patient, practice and unmodelled factors. Variation between practices was effectively explained, with residual variation between patients and within patients. The pWTD approach is useful for describing complex distributions of intervals, and may be more appropriate for inferring a gap than summarising truncated data.

**Plain language summary:** The time between one drug prescription and the next was calculated for the 20 most commonly prescribed drug classes in a database of older people attending primary care in Ireland. The timing varied drastically among drugs, due to differences in patients and practices. We analysed these variables using two approaches. The first looked at how factors such as age, sex, and healthcare cover relate to the time between prescriptions. The second described the overall pattern of prescription intervals as a combination of simpler underlying patterns.

**Key points:**

1. Prescription intervals are highly focussed around intervals of typical prescribed quantities, as expected, with modes of approximately 30, 60, and 90 days for all drug classes.
2. Heterogeneity in the prescription interval for specific drug classes was found to be mainly at the within-patient level. This may be due to prescription renewal behaviour changing over time for individuals.
3. Patient-specific covariates, such as age, sex and healthcare scheme cover had a similar effect on the prescription interval across the most frequent drug classes prescribed.
4. The parametric waiting time distribution, using a finite mixture model to account for unmodelled variation, was able to robustly explain the population average distribution of empirical intervals using a penalised likelihood procedure.

## Introduction

There is a growing volume of research evaluating the occurrence and effects of deprescribing; defined as a clinician-led intervention involving medication change with the goal of managing polypharmacy and improving outcomes. Routinely collected data, e.g. prescribing data from an electronic health record (EHR) is a useful data source for this research.(1) The robustness of such research is contingent on accurately determining deprescribing exposure,(2) and this is often done based on a static “gap” following the last prescription.(3) However, the reality of healthcare delivery and patient behaviour (i.e., irregular healthcare visits) can lead to variation in the timing of prescriptions which may impact on identifying cases of deprescribing using prescribing data. This variation, if not accounted for, may lead to exposure misclassification bias.(4,5) Another major factor that can contribute to exposure misclassification is non-adherence of prescribed or dispensed medication, however, adherence is unobservable in EHRs. In studies examining the effect of nonrandomised treatment strategies, the effect of this bias may be quantified using probabilistic bias analysis(6) however, such analyses require an estimate or distribution of the magnitude of misclassification, which is not available using standard gap-based definitions of exposure. Furthermore, in a recent review of methodology within deprescribing studies, 27% of studies did not clearly report how they defined the date of last medication exposure.(7) Where a fixed gap was used, it also varied among studies even within drug classes.(7) Thus, analysing prescription intervals (the time between prescriptions) may reduce reliance on a generalisable “gap” and could inform more accurate definitions of deprescribing.(8)

Estimating prescription intervals using a representative population is one approach to defining periods of continued treatment and discontinuation. (9) The prescription interval can be obtained by measuring the time from one prescription issue until the next renewal. In the present study, this is named the empirical interval method. This method is restricted to those who renew at least once. Modelling the empirical interval using a nonparametric approach provides a flexible summary of the distribution. When covariates are available, a more explanatory analysis can be performed by specifying a model for the intervals.

An alternative approach is to estimate the waiting time distribution (WTD).(9,10) The waiting time is the time from a single calendar index date (identical across the population) until the first observed prescription issue (not restricted to those who renew at least once). This method attempts to separate the incident from prevalent prescriptions. An estimate of the inter-arrival density (the probability density of renewal time for prevalent users) can be obtained from a parametric estimate of the WTD (pWTD), assuming the renewal process is stationary (i.e. the inter-arrival density is constant). The benefits over the empirical interval approach are the obviation of multiple renewals per patient and the potential to estimate the incidence of prescribing.(12) Using multiple random index dates has been shown to increase efficiency,(11) and may aggregate nonstationary effects.

This study aims to; (1) evaluate how prescription intervals (the time between successive prescriptions for a patient) in primary care vary across drug classes, patients, and GP practices, and (2) compare estimates of the interarrival density of renewals using the pWTD approach and empirical interval approach.

## Methods

A protocol was preregistered for this study on the Open Science Framework.(12) R version 4.5.0 was used for analysis.(13)

### Data source

This study incorporates a secondary analysis of prescription data from a cohort of Irish patients from 44 GP practices.(14) Data was extracted for patients aged 65 years and older for the period 2011-2020, containing details of WHO Anatomical Therapeutic Chemical (ATC) codes and dates of prescribing, as well as information regarding patients’ age, sex, number of prescription repeats and healthcare scheme cover.

### Data pre-processing

The 20 most frequently prescribed drug classes (excluding non-chronic drugs e.g., antibiotics) within the database were analysed. Prescriptions were classified into groups based on the fourth-level ATC code (i.e. chemical subgroup), with some classes combined if they spanned more than one fourth-level code e.g. benzodiazepines (see table S1 of the supporting information).

Covariates included age (continuous), sex (binary), health cover scheme (categorical: full public cover (General Medical Services scheme), partial public cover (Doctor Visit Card), and other/private cover: private cover and one other type), number of prescription repeats (integer), and anonymised GP practice and patient identifiers. The outcome was the interval between consecutive prescriptions. Intervals >540 days (∼18 months) were excluded as implausible within a single treatment course, with sensitivity analysis performed. When multiple prescriptions for the same drug occurred on the same date, prescription repeats were set to the maximum value. Observations with magnitude of Z-scores >3 for continuous/ordinal variables were excluded via listwise deletion, with sensitivity analyses assessing dependence. Continuous covariates were grand-mean centred.

### Analysis – empirical intervals

Prescription intervals were calculated and the marginal density estimated. Covariate associations were assessed using a multilevel linear model (MLM) with restricted maximum likelihood (REML) in *lme4*.(15) Two models were fitted, one including random intercepts only (unconditional mean model) as recommended,(16) and one including random intercepts and fixed covariate effects (full model). The full model was specified as

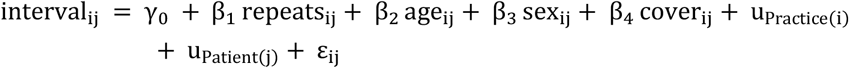

and the mean model as

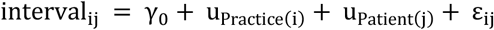

Where γ_0_ is the grand mean intercept, β_i_ the coefficients, repeats_ij_ is the number of prescription repeats, cover_ij_ is the healthcare cover type, u_Practice(I)_ is the GP practice random effect, u_Patient(j)_ the patient random effect and ε_ij_ is the residual term.

### Analysis – waiting time

The WTD in this context is the distribution of time from an index date D to the next prescription renewal R_i_ for all patients.(17) The WTD density *h(R*_*i*_*-D)* can be modelled as

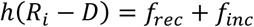

where *f*_*rec*_ is a forward recurrence density (FRD) and *f*_*inc*_ is an incidence density (see supporting information for details). When the renewal process is in equilibrium (the time origin of the process is in the distant past), *f*_*rec*_ is a function of *f*_*IAD*_ only(18)

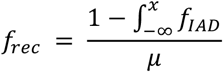

Where µ is the mean of *f*_*IAD*_. Thus, a parametric form can be assumed for *f*_*IAD*_ and *f*_*inc*_, and *h*(*R*_*i*_ − *D*) can be fit to the data. *f*_*IAD*_ can then be obtained through plug-in. This method has previously been implemented using the maximum likelihood estimation (MLE) procedure assuming a two-component mixture of recurrence densities and a single uniform incidence density.(19) While a global MLE does not exist for a finite mixture of location-scale distributions(20), obtaining estimates from the local maxima of the likelihood are informative when convergence is achieved.(21) In this work, the model was extended to an arbitrary but finite number (*g*) of recurrence densities. This was due to the added flexibility of modelling a larger number of clusters, due to package sizes or typical prescription durations. (15) Ten random index dates were used and *g* was varied from 1 to 5. All *f*_*IAD,i*_ were chosen to be from the lognormal family due to its non-negative support. The Bayesian Information Criterion (BIC) was used to find the optimal *g* value.(22) and the Akaike Information Criterion (AIC) was also reported. The BIC is given as

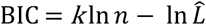

Where *k* is the number of parameters estimated by the model, *n* is the number of samples and 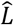 is the maximised likelihood. After obtaining the optimal *g* (best of 5 runs), a separate model was fit again to obtain the WTD parameters. The code is on Zenodo.(23)

## Results

Overall, 38,154 patients who were prescribed one of the relevant drug classes were included, with a mean age of 78 years and 43% were male (see Table 1). Across the included drug classes, 2.65 million renewals occurred, with a median of 3 prescription repeats (IQR 1-3). Statins, proton pump inhibitors and paracetamol were the three most common drug classes prescribed.

**Table 1.** Data characteristics after pre-processing.

| Characteristic | N(%)<sup>1</sup> |
| --- | --- |
| Patient characteristics (n=38.154) |  |
| Age, mean (SD) | 78 (9) |
| **Sex** |  |
| Female | 21,628 (57%) |
| Male | 16,526 (43%) |
| **Health cover type** |  |
| Partial public cover (Doctor Visit Card) | 4,111 (11%) |
| Full public cover (General Medical Services scheme) | 26,574 (70%) |
| Other/private | 7,469 (20%) |
| Prescription characteristics (n=2,652,118) |  |
| Prescription repeats, median (IQR) | 3.00 (1.00, 3.00) |
| **Drug** |  |
| Statins | 365,633 (14%) |
| Proton pump inhibitors | 287,605 (11%) |
| Paracetamol | 219,542 (8.3%) |
| Angiotensin receptor blockers +/- diuretics | 207,211 (7.8%) |
| Beta blockers | 199,478 (7.5%) |
| Calcium +/- vitamin D | 171,724 (6.5%) |
| Antiplatelets (incl. low dose aspirin) | 162,441 (6.1%) |
| Benzodiazepines and associated drugs | 151,175 (5.7%) |
| ACE inhibitors +/- diuretics | 122,666 (4.6%) |
| DHP calcium channel blockers +/- diuretics | 113,516 (4.3%) |
| Laxatives | 103,304 (3.9%) |
| SSRIs and SNRIs | 103,020 (3.9%) |
| Inhaled adrenergics | 80,245 (3.0%) |
| Inhaled adrenergics with corticosteroids and/or anticholinergics | 75,062 (2.8%) |
| Thyroid hormones and derivatives | 66,004 (2.5%) |
| NSAIDs | 54,236 (2.0%) |
| Opioids | 50,436 (1.9%) |
| Oral anticoagulants | 46,187 (1.7%) |
| Gabapentinoids | 38,886 (1.5%) |
| Other antidepressants | 33,747 (1.3%) |

<sup>1</sup>Unless otherwise stated

### Empirical distribution and multi-level modelling

The number of prescription interval pairs per drug class varied from 365,633 (statins) to 33,747 (other antidepressants). The marginal inter-arrival density (IAD) is shown in Figure 1 (a). The graph shows a multimodal structure with modes spaced by approximately one month (as expected due to the typical intended prescription durations). Beyond 120 days there were low numbers of renewals across all drug classes, potentially due to different treatment episodes. The analysis was also stratified by the number of prescription repeats, and multi-modality persisted (see Figure S2). The marginal distribution is shown in Figure S3.

**Figure 1.**
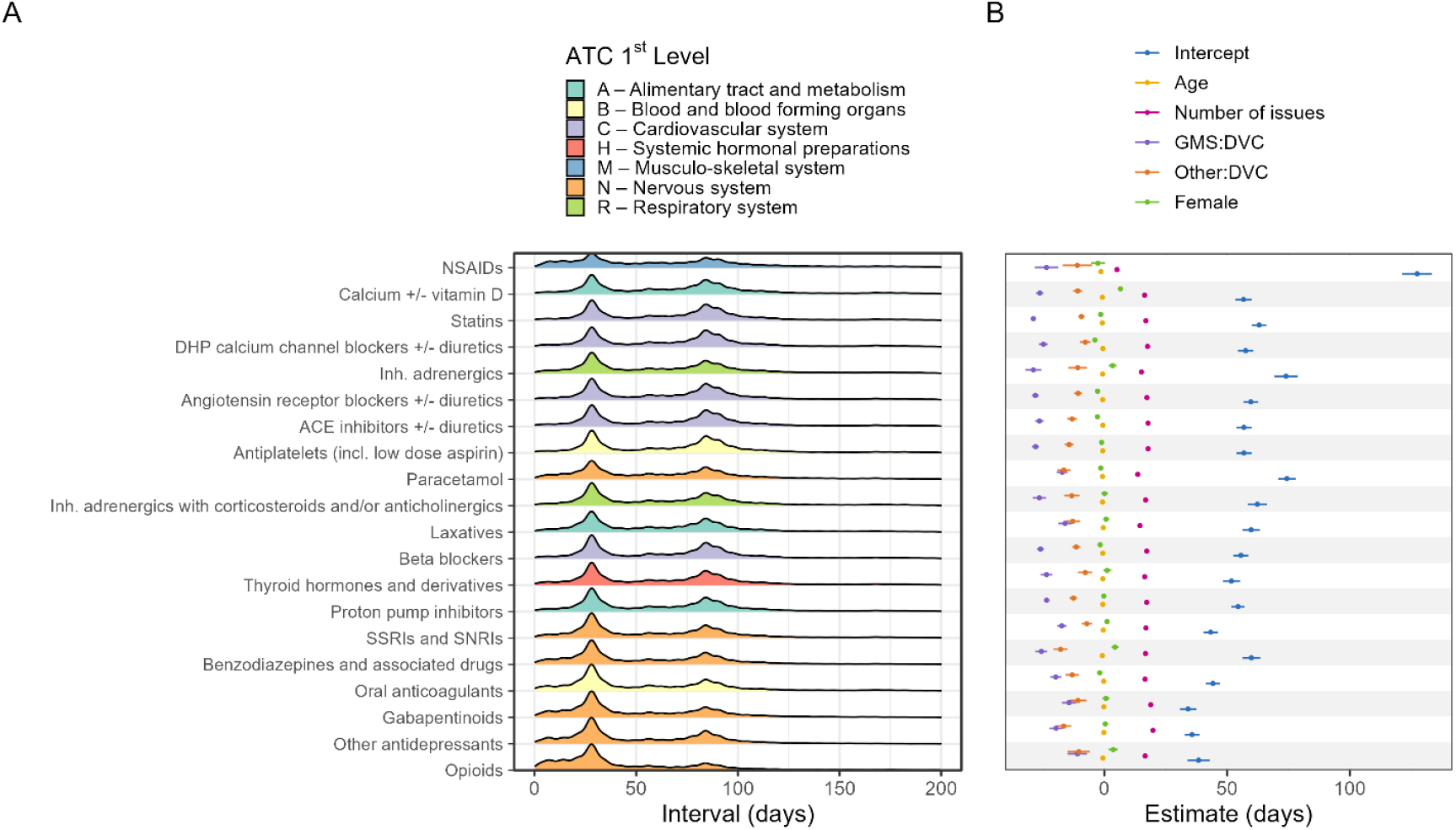
(a) Kernel density estimate of the inter-arrival distribution for all drug classes. The drugs are ordered by mean interval descending. (b) Coefficients and population-level intercept for the multi-level model. GMS: full public cover, DVC: partial public cover, Other: private cover and one other type.

Figure 1 (b) shows the fixed effects coefficients and grand mean intercept (across-level mean interval when covariates are equal to the reference value) with their 95% confidence intervals obtained from the full model. The grand mean intercept varied substantially, from 34.0 days (gabapentinoids) to 127.4 days (NSAIDs), as seen in Figure 1 (b). For 10 drug classes the intercept was in the range of 50-60 days. The number of prescription repeats was associated with an increase in the prescription interval by 5.0 days (NSAIDs) to 19.7 days (Other antidepressants), per additional prescription issue. A patient being female was associated with a -3.9 day (dihydropyridine calcium channel blockers +/-diuretics) to 6.5 day (Calcium +/-vitamin D) change relative to being male. A one-year difference in age was associated with a -1.4 day (NSAIDs) to -0.2 day (Other antidepressants) change in the interval. Finally, full public cover was associated with a -29.0 day (Inhaled adrenergics) to - 11.0 day (Opioids) change relative to partial public cover patients, while those with other/private cover had an associated -17.9 day (Benzodiazepines and associated drugs) to - 7.1 day (SSRI and SNRIs) change relative to partial public cover scheme patients.

The variance at each level for the null and full models is shown in Figure 2. The distribution of the random intercepts is shown in Figure S3. The MLM results without outlier removal and after truncating at a shorter gap of 270 days are also shown in Figure S5 and Figure S6 and S7.

**Figure 2.**
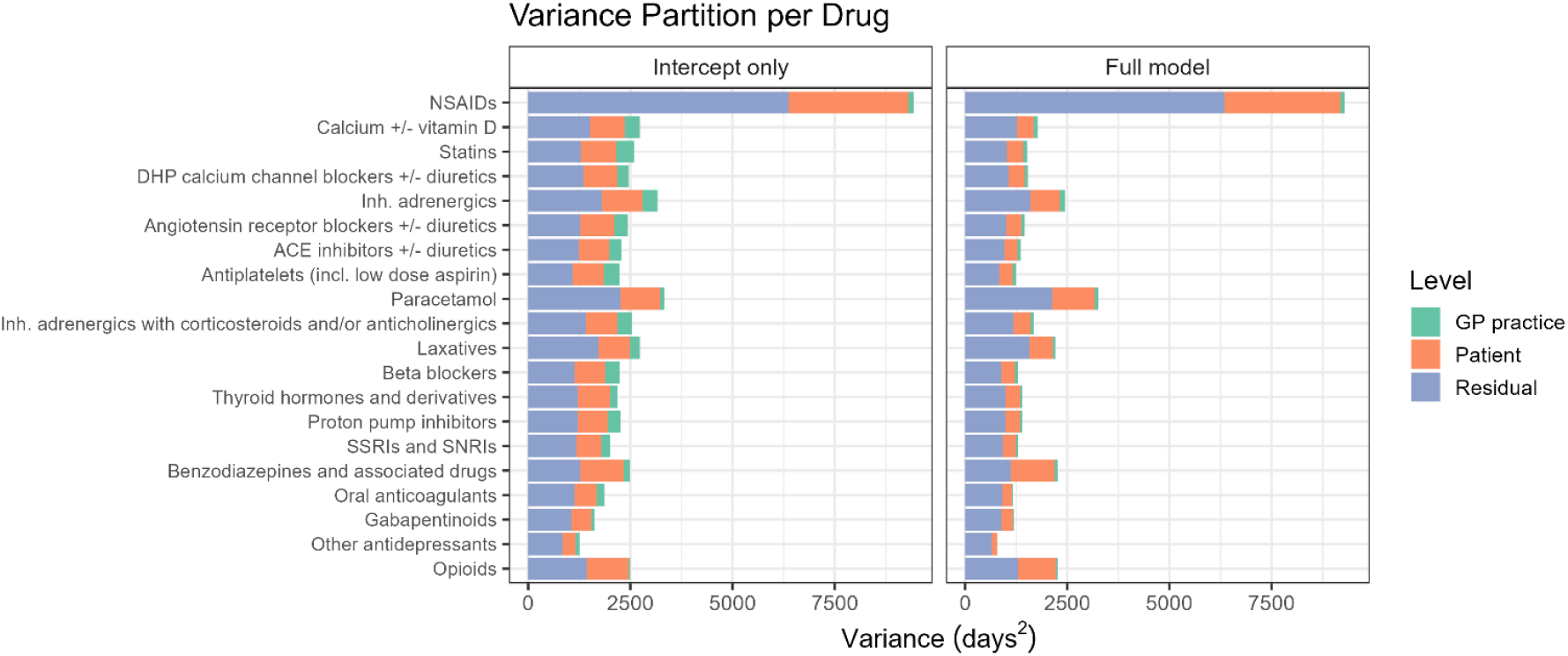
Variance of each model by group level.

### Parametric waiting time distribution

The results of the model selection procedure are shown in Figure *3* (a), plotting the BIC and AIC versus the number of forward recurrence density (FRD) components. This showed that 3 was the optimal value of *g* for 10 of the classes. The results of the fitting procedure are shown in Figure *3* (b) using QQ plots. The median and 80^th^ percentile for the empirical and modelled inter-arrival density is shown in Table 2, which generally showed good agreement between empirical and fitted densities over the chosen quantiles, with NSAIDs, Angiotensin receptor blockers +/-diuretics, Antiplatelets (incl. low dose aspirin), Paracetamol, Beta blockers, SSRIs and SNRIs and Oral anticoagulants classes being an exception.

**Table 2.** Empirical and fitted inter-arrival density values at the 0.5 and 0.8-quantile. Confidence intervals are pointwise, based on 20 bootstrap samples.

| Quantile | 0.5 |  | 0.8 |  |
| --- | --- | --- | --- | --- |
| Drug | Empirical | Fitted (95% CI) | Empirical | Fitted (95% CI) |
| NSAIDs | 77 | 71.3 (67.4–75) | 149 | 112.1 (107.2–120.8) |
| Calcium +/- vitamin D | 76 | 78.1 (76.2–79.4) | 103 | 102.1 (98.9–105) |
| Statins | 77 | 74.6 (73.2–75.6) | 100 | 99.7 (98.7–100.4) |
| DHP calcium channel blockers +/- diuretics | 77 | 74 (71.6–76.5) | 100 | 100.1 (98.1–102.1) |
| Inh. adrenergics | 69 | 64.2 (58.1–72.1) | 100 | 98.7 (96.2–101.8) |
| Angiotensin receptor blockers +/- diuretics | 76 | 71.6 (70–74.1) | 98 | 95 (93.8–95.9) |
| ACE inhibitors +/- diuretics | 75 | 72.7 (71.4–73.8) | 97 | 96.3 (95.1–97.7) |
| Antiplatelets (incl. low dose aspirin) | 76 | 73.5 (71.6–75.1) | 97 | 99.9 (98.5–101.2) |
| Paracetamol | 63 | 68.4 (65.4–70.5) | 98 | 106.2 (103.4–110.7) |
| Inhaled adrenergics with corticosteroids and/or anticholinergics | 67 | 60.7 (55.8–66.2) | 98 | 99.9 (97.6–102.2) |
| Laxatives | 66 | 56.4 (53.9–59.5) | 96 | 98.2 (95.8–101.4) |
| Beta blockers | 71 | 75.4 (73.5–77.5) | 95 | 98.6 (97.2–99.7) |
| Thyroid hormones and derivatives | 67 | 68.9 (65.2–72.2) | 95 | 97.5 (95.2–99.6) |
| Proton pump inhibitors | 65 | 67.9 (66.7–69.2) | 94 | 96.6 (94.7–99) |
| SSRIs and SNRIs | 56 | 62.1 (57.9–66) | 91 | 96.4 (94–98.1) |
| Benzodiazepines and associated drugs | 50 | 49.4 (46.8–52.1) | 90 | 94.8 (93.2–96.4) |
| Oral anticoagulants | 46 | 35.4 (35.2–36.8) | 88 | 93.4 (93.3–94.4) |
| Gabapentinoids | 39 | 47.7 (44.3–49.3) | 86 | 86.6 (84.2–88.8) |
| Other antidepressants | 42 | 46.8 (43.9–50.5) | 85 | 87.8 (86.2–90.5) |
| Opioids | 31 | 35.4 (34–37.5) | 83 | 82.6 (78.8–86.6) |

## Discussion

The most frequent intervals for all drug classes were centred around 30 and 90 days, consistent with a similar study in England.(24) There was also markedly similar shapes in the marginal distributions between drug classes. The extended intervals in the right tail of the interval distribution may be due to an increased quantity of medication being prescribed at the previous visit, for example when anticipating travel, or different treatment courses for some drugs or prescriptions given during hospitalisation. There was a distinct probability mass around <25 days for NSAIDS, other antidepressants, benzodiazepines, gabapentinoids and opioids. These may be due to extending short courses of treatment (e.g. opioid treatment for one week following injury being reissued due to persisting pain), or smaller quantities being prescribed due to risk of misuse. The fixed effect coefficients were largely unchanged for the smaller choice of interval truncation (see Figure S6 and S7) but the intercepts and variance were reduced. The residual variance of the NSAID class was also more comparable with the rest of the drug classes for this case, suggesting that intervals beyond 270 days significantly contributed to the original residual variance for this class (likely repeated acute use). The MLM exhibited heteroskedasticity and nonlinear residuals and the standard errors of the fixed effects were expected to be biased as a result of the misspecification due to omitted terms and/or parametric assumptions being violated. Therefore, no hypothesis tests were conducted.

Figure 2 shows that the largest contributor of variance in the prescription interval is within patients, with the between-patient level accounting for the second largest source of variance. GP practice level was the lowest source for all drug classes. High within-patient variation in the interval may result in misclassification of exposure and large numbers of identified intercurrent events such as restarting. The change in the total variance when patient covariates were included, as shown in Figure 2 and tabulated in Table S8 shows that the covariates explain some of the variation, however the change in total variance was negligible for some drug classes such as paracetamol and NSAIDs. This could be due to latent unmeasured variables at patient or practice level. The GP practice-level variance decreased for all drug classes compared to the null model. This was likely due to a different distribution of characteristics of patients between practices, which may be associated with the prescription interval. The multimodal nature of the marginal IAD was partly explained by the number of prescription repeats (i.e. individual repeats per prescription). The residual variance (shown in Figure S2) may be due to deviations in renewal behaviour, such as re-issues before the end of supply. This means there was unmodelled heterogeneity, which can be modelled using the WTD approach because a mixture of distributions are possible.

The model selection results for the WTD show that the optimal *g* (minimising BIC) was between 2 and 5, with 10 of the drug classes having an optimal of 3. The number of components may correspond with latent subpopulations that share a common characteristic, such as prescribed duration of the drug, or from systematic data differences e.g. software representation or prescriber behaviour. For all but one class, the BIC curve was convex, highlighting that the range of *g* was appropriate. Since the likelihood surface may be non-convex in a mixture model, the choice of starting values for the maximum likelihood optimisation may have resulted in a local minimum being found.(25) Therefore, increasing the number of replications for the optimisation may have resulted in a different value of *g* for that drug class.

The WTD and empirical distribution had good agreement for the majority of drug classes as seen in Figure 3 (b). Table 2 also shows very close agreement between the empirical and fitted median IAD. There were two noticeable regions of lack-of-fit. Firstly, in the right tail there was systematic over-estimation of the IAD for certain classes e.g., Antiplatelets (incl. low dose aspirin), Paracetamol, Beta blockers and Oral anticoagulants. This was also apparent for the 80^th^ percentile in Table 2. This may be due to the lack of data at the longer intervals. Under-estimation occurred for NSAIDS and laxatives, possibly due to the artificial truncation of intervals in the pre-processing stage, which necessarily biases the waiting time downward. Since this occurs in the upper quantiles of the empirical distribution, the effect of this should not severely affect the interpretation of model parameters. The other observed feature in the QQ plots was a non-smooth trend at approximately 30 days in the Calcium +/-vitamin D, Statins, DHP calcium channel blockers +/-diuretics, Paracetamol, Beta blockers, Proton pump inhibitors Oral anticoagulants and Gabapentinoids classes. This feature was likely due to distributional assumptions being violated. Figure 1 (a) shows significant probability mass in the range of zero to 30 days that would not be well described by a lognormal distribution. Other distributions such as a Weibull distribution could be explored, which may produce better fit in this region. When the MLM was fit without outlier removal, the precision of the fixed effects estimates decreased due to larger variance in the response (see Figure S5). The intercept also had substantial variation compared to the pre-processed data. However, the fixed effects did not differ drastically from the pre-processed results.

**Figure 3.**
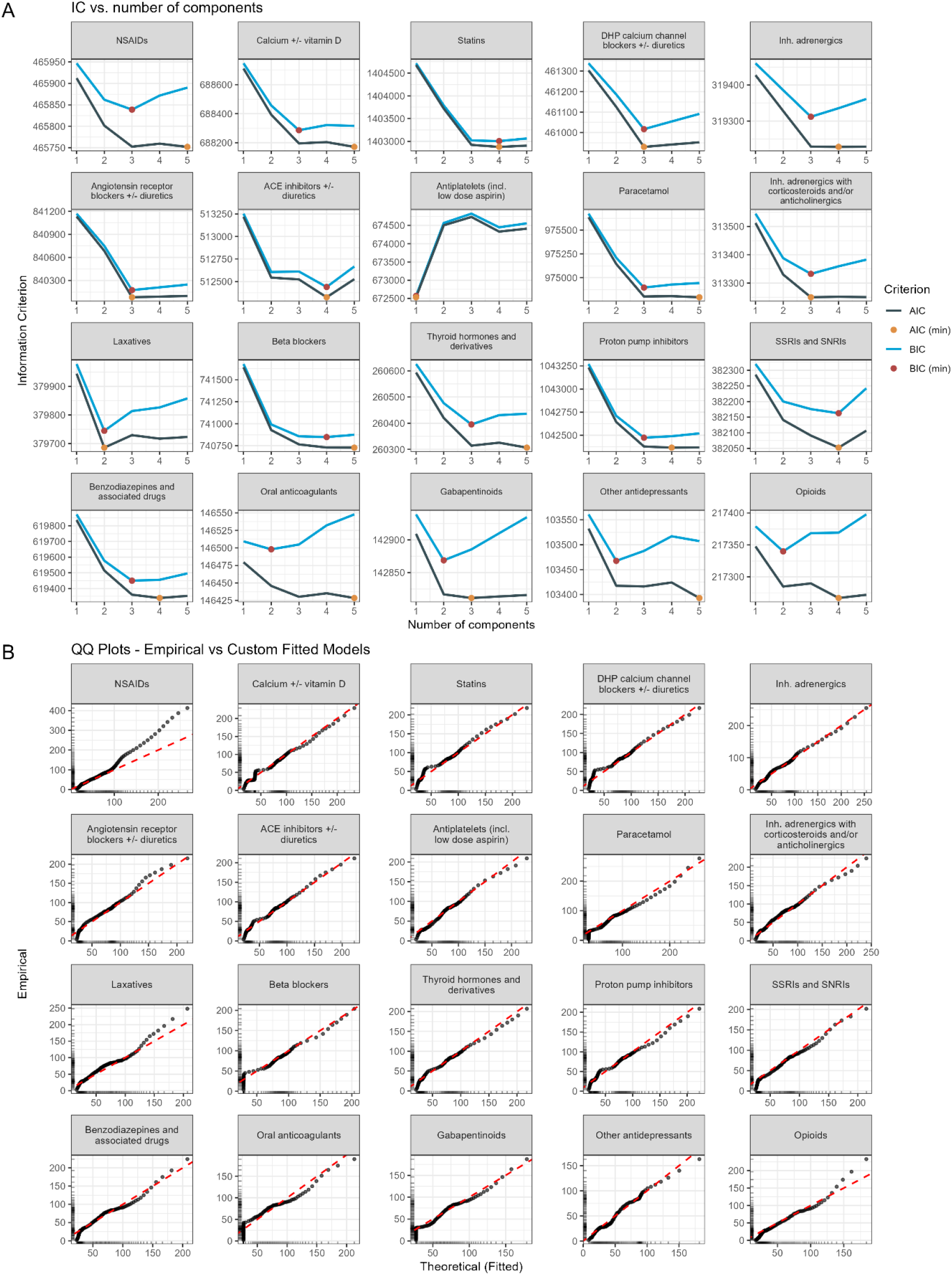
(a) AIC and BIC with their minima labelled vs. the number of mixture components stratified by drug class, (b) QQ plots of the empirical vs model-based distribution stratified by drug class.

Conditioning on the presence of at least two prescriptions within a prespecified time window was done in order to limit the analysis to prevalent users. A limitation is that termination of the renewal process or administrative censoring was not accounted for. The process may terminate due to a variety of reasons, such as deprescribing, death, patient-led decisions (non-adherence or change of provider), or external factors. Estimating the time when termination occurs in an observational database is difficult, since no information is typically recorded when this occurs.(26) When analysing prevalent users however, the IAD can be used to extrapolate a “stopping time” estimate using a decision rule. When applied to chronic medications there is a risk of underestimating the true stopping time due to stockpiling and non-adherence. Thus, a sensitivity analysis is recommended.(27) A decision rule based on the IAD for defining exposure is a crude method that should be used when no other patient- or prescriber-related information is available.

## Conclusions

This study analysed prescription intervals for the most frequently prescribed drugs in a GP database of older adults in Ireland. The interval was highly variable within drug classes and the variation was mostly due to sporadic patient-level renewal times that were not adequately explained by the measured covariates. Thus, observational studies may benefit from using data-informed static gaps when prescription-free periods are used to define exposure, as this may improve classification accuracy. Gaps that are excessively long can truncate available follow-up time and may render outcomes occurring close to exposure unidentifiable. Across all drug classes, the fixed effects of patient/prescription characteristics had consistent direction and magnitude of association with the renewal time, suggesting that the covariates exerted a similar influence on the interval regardless of the type of drug prescribed. Consequently, dynamic gaps are unlikely to provide additional benefit.

The parametric waiting time distribution is useful for characterising prescription intervals without requiring conditioning on prevalent users. Incorporating mixtures of distributions allows the modelling of the complex interval distribution. Prevalent and incident-user partitioning is possible under certain assumptions. When implementing a population-based decision rule to define medication discontinuation, such as one derived from the marginal parametric WTD, substantial prediction error may remain at the individual level. However, compared with simpler single-distribution or conditional approaches, mixture models may provide more accurate estimation of the population renewal-time distribution and therefore provide better-calibrated static discontinuation measures, as they explicitly account for heterogeneity in renewal behaviour.

## Data Availability

All data are anonymised electronic health records and so are unable to be shared.

https://doi.org/10.5281/zenodo.20411184

