## Supplementary information for "Prescription intervals of medications for chronic use: a cohort study"

### Supporting information

*Table S 1 ATC groupings*

| ATC code | Grouping |
|---|---|
| A02BC | Proton pump inhibitors |
| A06A | Laxatives |
| A12AX | Calcium +/- vitamin D |
| A12AA | |
| A11CC04 | |
| A11CC05 | |
| A11CC03 | |
| B01AC04 | Antiplatelets (incl. low dose aspirin) |
| B01AC24 | |
| B01AC22 | |
| B01AC06 | |
| B01AA03 | Oral anticoagulants |
| B01AB10 | |
| B01AB04 | |
| B01AB05 | |
| B01AE07 | |
| B01AF01 | |
| B01AF02 | |
| B01AF03 | |
| B01AB01 | |
| C07AB | Beta blockers |
| C08CA02 | DHP calcium channel blockers +/- diuretics |
| C08CA05 | |

| ATC code | Grouping |
| --- | --- |
| C08CA13 | |
| C08GA02 | |
| C08CA01 | |
| C09CA04 | Angiotensin receptor blockers +/- diuretics |
| C09CA06 | |
| C09DA03 | |
| C09CA07 | |
| C09CA08 | |
| C09CA03 | |
| C09CA01 | |
| C09DA07 | |
| C09DA04 | |
| C09DA01 | |
| C09BA04 | ACE inhibitors +/- diuretics |
| C09AA02 | |
| C09AA04 | |
| C09AA05 | |
| C10AA | Statins |
| H03AA | Thyroid hormones and derivatives |
| M01AE52 | NSAIDs |
| M01AE51 | |
| M01AH02 | |
| M01AH01 | |
| M01AG01 | |
| M01AE02 | |
| M01AE01 | |
| M01AB05 | |
| N02BE51 | Paracetamol |
| N02BE01 | |
| N02AJ13 | Opioids |
| N02AX02 | |
| N02AA01 | |
| N02AA05 | |
| N02AA08 | |
| N02AA56 | |
| N02AB03 | |
| N02AA55 | |
| N02AJ06 | |
| N02AA59 | |
| R03DA03 | |
| R05DA04 | |
| N02AE01 | |
| N03AX12 | Gabapentinoids |

| ATC code | Grouping |
| --- | --- |
| N03AX16 | |
| N06AX01 | Other antidepressants |
| N06AX02 | |
| N06AX03 | |
| N06AX04 | |
| N06AX05 | |
| N06AX06 | |
| N06AX07 | |
| N06AX08 | |
| N06AX09 | |
| N06AX10 | |
| N06AX11 | |
| N06AX12 | |
| N06AX13 | |
| N06AX14 | |
| N06AX15 | |
| N06AX17 | |
| N06AX18 | |
| N06AX19 | |
| N06AX22 | |
| N06AX23 | |
| N06AX24 | |
| N06AX25 | |
| N06AX26 | |
| N06AX27 | |
| N06AX28 | |
| N06AX29 | |
| N06AX31 | |
| N06AX62 | |
| N06AB03 | |
| N06AB04 | SSRIs and SNRIs |
| N06AB05 | |
| N06AX21 | |
| N06AX16 | |
| N06AB10 | |
| N06AB06 | |
| N05BA01 | |
| N05BA12 | Benzodiazepines and associated drugs |
| N05CD07 | |
| N05CD08 | |
| N05CF02 | |
| N05BA08 | |
| N05BA02 | |

| ATC code | Grouping |
|---|---|
| N05BA04 | |
| N03AE01 | |
| N05BA09 | |
| N05CF01 | |
| N05BA06 | |
| R03AC | |
| R03AK07 | Inhaled adrenergics |
| R03AK06 | Inhaled adrenergics with corticosteroids and/or anticholinergics |
| R03BB | |
| R03AK11 | |
| R03AK08 | |

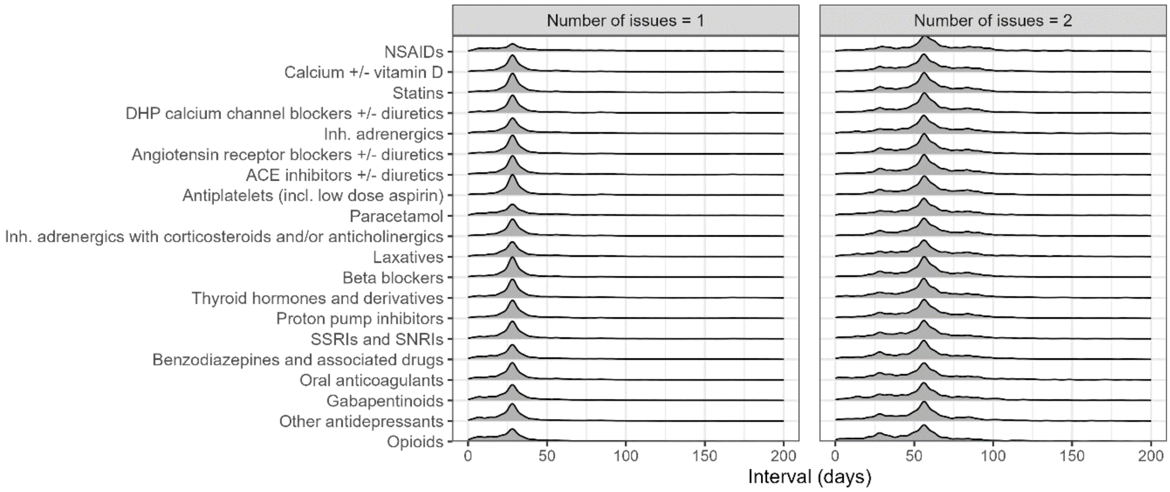

*Figure S 2 Marginal empirical interval stratified by drug class and number of prescription repeats.*

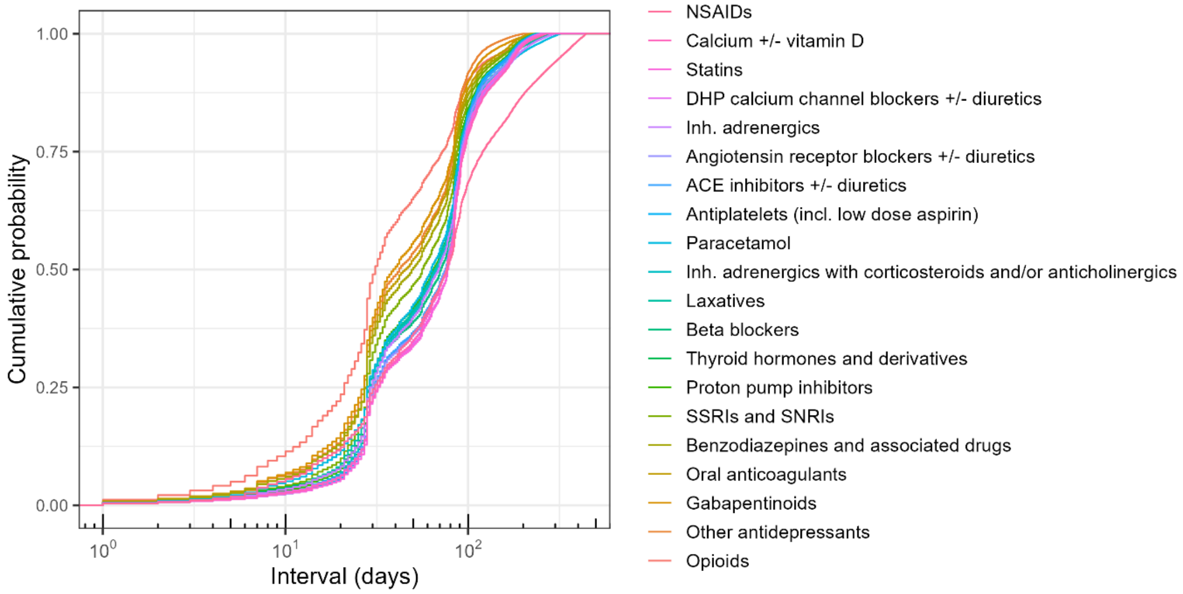

*Figure S 3 Marginal empirical distribution functions for each drug class*

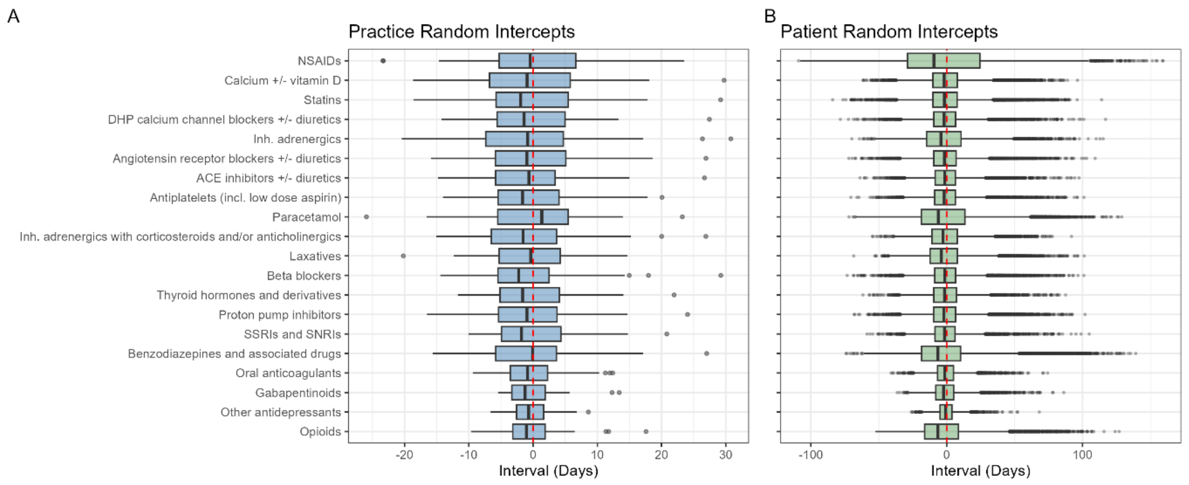

*Figure S 4 (a) Boxplot of the distribution of practice-level random intercepts (b) boxplot of the distribution of patient-level random intercepts.*

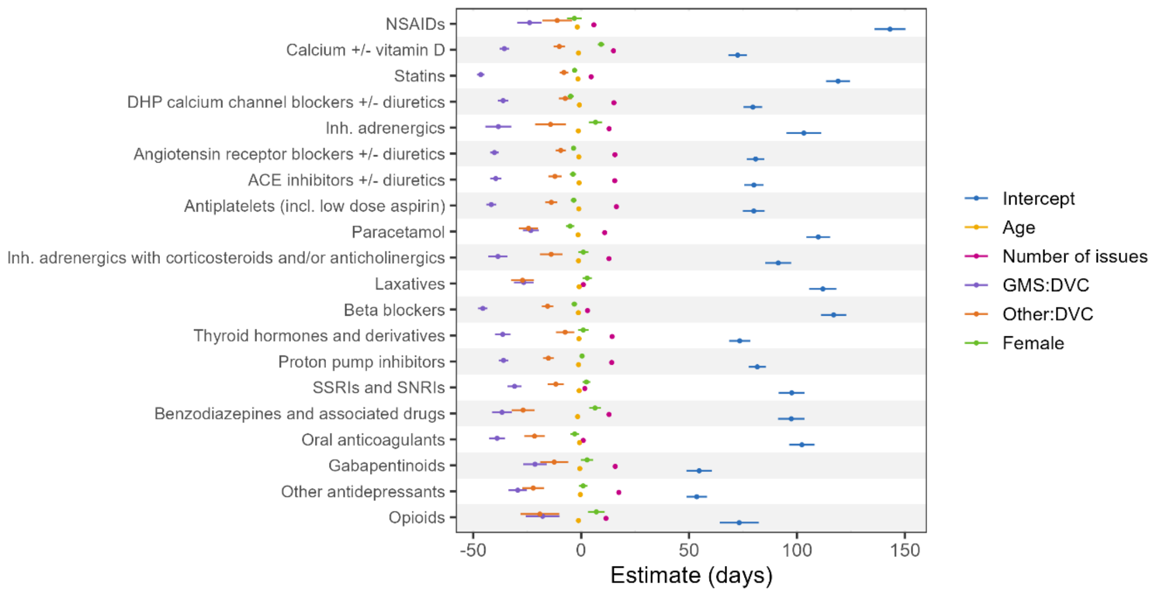

*Figure S 5 Fixed effects from the multi-level model without covariate outlier removal.*

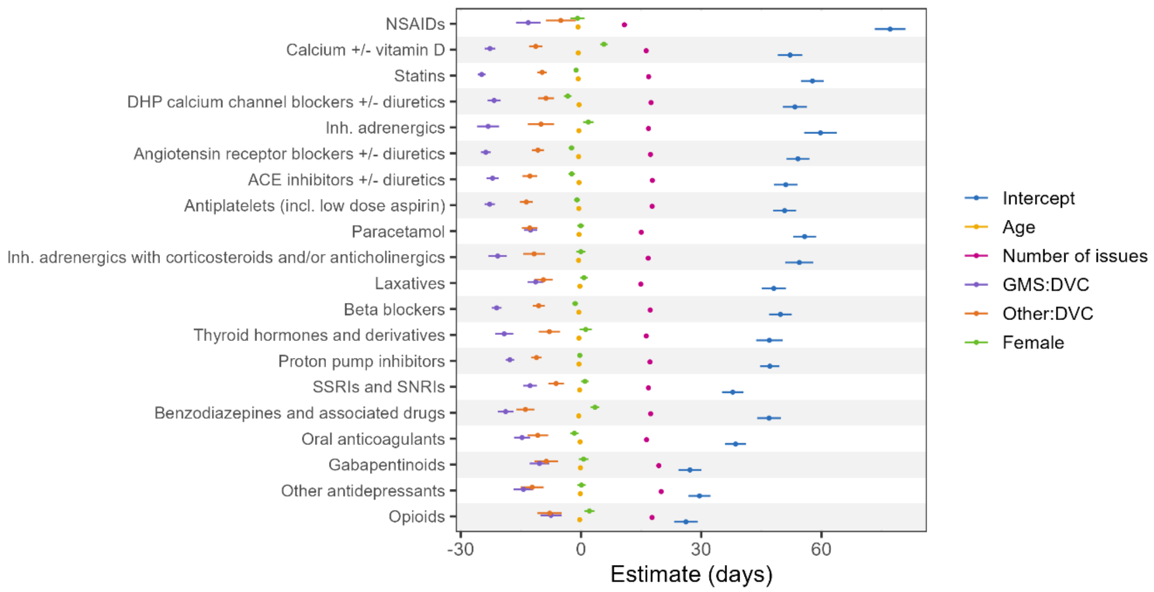

*Figure S 6 Fixed effects from the multi-level model with intervals truncated at 270 days.*

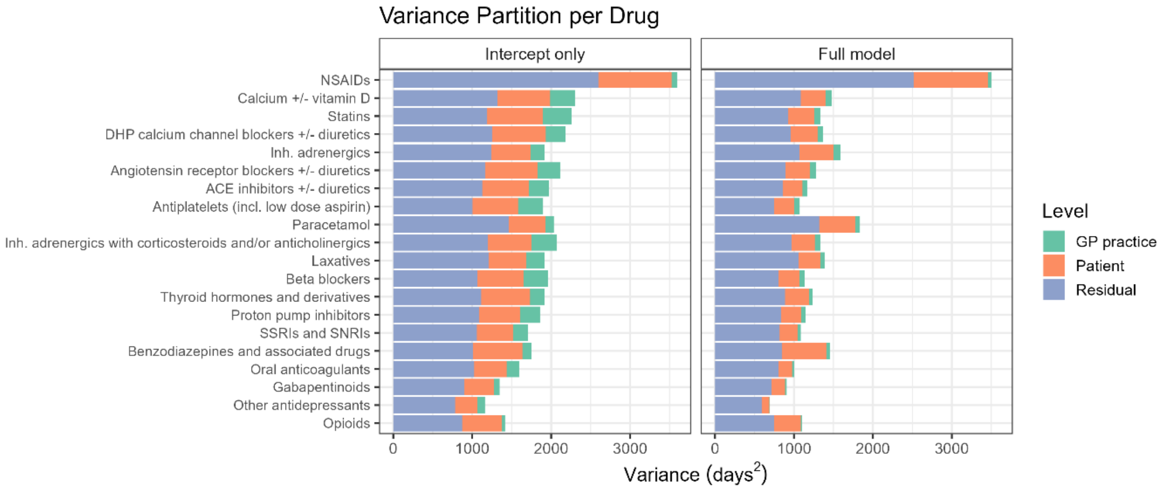

*Figure S 7 Variance by level with intervals truncated at 270 days.*

*Table S 8 Model variance by drug class, model and level.*

| Drug | Intercept only | | | Full model | | |
|---|---|---|---|---|---|---|
| | Residual | GP level | Patient level | Residual | GP level | Patient level |
| NSAIDs | 6,381.4 | 2,923.2 | 121.2 | 6,346.4 | 2,834.5 | 111.9 |
| Calcium +/- vitamin D | 1,506.6 | 849.5 | 380.8 | 1,273.3 | 409.3 | 88.9 |
| Statins | 1,292.4 | 874.6 | 429.7 | 1,026.2 | 402.3 | 88.0 |
| DHP calcium channel blockers +/- diuretics | 1,357.2 | 820.7 | 276.3 | 1,059.2 | 406.7 | 72.3 |
| Inhaled adrenergics | 1,796.8 | 1,000.3 | 367.2 | 1,592.9 | 721.2 | 123.9 |
| Angiotensin receptor blockers +/- diuretics | 1,272.1 | 837.2 | 332.1 | 999.3 | 385.4 | 80.9 |
| ACE inhibitors +/- diuretics | 1,233.8 | 753.4 | 291.8 | 960.6 | 325.2 | 74.9 |
| Antiplatelets (incl. low dose aspirin) | 1,092.0 | 756.4 | 387.7 | 832.9 | 331.5 | 81.2 |
| Paracetamol | 2,264.1 | 958.9 | 114.0 | 2,132.0 | 1,042.1 | 83.5 |
| Inhaled adrenergics with corticosteroids and/or anticholinergics | 1,410.0 | 767.5 | 353.6 | 1,181.0 | 421.5 | 82.5 |
| Laxatives | 1,719.9 | 769.0 | 239.7 | 1,569.2 | 580.3 | 58.9 |
| Beta blockers | 1,148.1 | 737.2 | 357.3 | 880.0 | 334.5 | 79.2 |
| Thyroid hormones and derivatives | 1,210.7 | 783.9 | 196.0 | 978.2 | 365.9 | 55.8 |
| Proton pump inhibitors | 1,225.6 | 713.5 | 312.7 | 977.9 | 357.5 | 61.3 |
| SSRIs and SNRIs | 1,181.7 | 608.3 | 212.4 | 931.6 | 311.7 | 52.0 |
| Benzodiazepines and associated drugs | 1,280.0 | 1,061.1 | 151.0 | 1,118.8 | 1,060.5 | 77.0 |
| Oral anticoagulants | 1,139.6 | 535.7 | 196.3 | 908.7 | 227.4 | 32.7 |
| Gabapentinoids | 1,062.1 | 476.9 | 77.8 | 881.1 | 292.5 | 23.8 |
| Other antidepressants | 842.1 | 314.2 | 98.4 | 650.0 | 128.4 | 14.8 |
| Opioids | 1,427.8 | 1,023.8 | 53.4 | 1,308.5 | 924.5 | 36.3 |

#### WTD model details

The density of the WTD is usually assumed to contain a prevalent component (for renewals of prescriptions) and an incident component (for first time prescriptions). The prevalent component corresponds to a forward (or backward, depending on whether the next or last prescription is used) recurrence density $f_{rec}$ which is a length-biased estimate of the interarrival probability density. The incidence density $f_{inc}$ gives the distribution of new prescriptions and is typically assumed to be constant over the period of interest.

The WTD is parameterised as

$$h(R_i - D) = w\left(\sum_{i=1}^{g} p_i \frac{1 - \int_{-\infty}^{x} f_{IAD,i}}{\mu_i}\right) + \frac{(1-w)}{\Delta}$$

where $w$ is the proportion of prevalent patients, $p_i$ is the prior probability of the waiting time being generated from component $f_{IAD,i}$ and $\Delta$ is the width of the time interval of interest. The waiting time distribution was estimated for each drug class using multiple random index dates. Each index was randomly sampled from a uniform distribution over ($t_0$, $t_0+\Delta$), where $t_0$ is the earliest prescription renewal date and $\Delta$ was chosen as 365 days. Using multiple random index dates improves the estimate of the FRD by increasing the number of available samples for the optimization. While using random index dates will necessarily induce dependence in the waiting times among samples, the marginal distribution will be unaffected. However, the inclusion of covariates in the WTD model will require consideration as the lack of independence may render the parameter estimates to be biased, and potentially induce further problems in downstream analyses incorporating these estimates (such as biased effect estimates).

The optimal parameters returned by the MLE procedure are known to be dependent on initial conditions due to the presence of nonconvex likelihood surfaces, so we used the best fitting model (in terms of likelihood) from 5 independent runs. The MLE procedure used the *optim* function with the L-BFGS-B optimization method, a quasi-Newton method incorporating box constraints, and a maximum number of 50 iterations was used. We set a lower bound of 0.05 on $\sigma$ (the standard deviation on the exponential scale) of each interarrival density to prevent spurious fitting to sparse regions, and initialised the distributional parameters using a Gaussian finite mixture model (MClust) of the logarithm of the waiting times. We also regularised $\sigma$, w and p using the exponential, softmax and logistic functions respectively. All other parameters were defaults.
